# Satisfaction and willingness-to-pay for water services in rural northern Ghana: Do we need intermediary steps toward safely managed services?

**DOI:** 10.1101/2025.11.10.25339759

**Authors:** John Trimmer, Joyce Kisiangani, Bessy Ewoenam Odame-Boafo, Lisa Appavou, Chloé Poulin, Dominic Osei, Caroline Delaire, Valerie Bauza

## Abstract

Universal access to safely managed water, defined as water from an improved source that is accessible on premises, available when needed, and free from contamination, remains far off in many low-resource settings. Progress toward safely managed services may include intermediate steps focused on improving some service attributes over others. However, knowledge surrounding households’ service preferences remains limited. This study explored satisfaction with existing water service characteristics and willingness-to-pay for hypothetical improvements in water supply. We conducted surveys with 1,748 households among 120 rural communities in northern Ghana. While few households (<2%) had safely managed water services, approximately half (52%) had at least basic access (i.e., an improved source where collection time is within 30 minutes). Satisfaction with existing water services was associated with source type, accessibility, availability, and perceived safety. For example, respondents who always had drinking water available in sufficient quantities had almost three times the odds of being satisfied (odds ratio [OR]: 2.66, 95% CI: 1.94-3.66), similar to respondents able to collect water within 30 minutes (OR: 3.09, 95% CI: 2.28-4.19) and those with on-premises water access (OR: 2.90, 95% CI: 1.26-6.66). Accessibility and water quality were particularly high priorities. Using discrete choice experiments, we estimated participants were willing to pay 3.6-4.9 times more for on-premises piped connections (compared with handpumps 30 minutes away) and chlorination (compared with no treatment). To a lesser extent, households were willing to pay for improved reliability, increased availability, and shorter collection times (10 vs. 30 minutes). Accordingly, we considered the possibility of introducing an intermediate service level: “proximate access”, defined as using an improved source where water is collected within 10 minutes, available when needed, and (possibly) free from contamination. Such a category may provide feasible intermediate improvements to advance progress toward more highly valued safely managed services in rural, low-resource settings.

## 1. Introduction

Safely managed water services, the gold standard promoted by the Sustainable Development Goals (SDGs), require water from an improved source that meets three criteria for users: it is accessible on premises, available when needed, and free from contamination [1]. Worldwide, 38% of rural residents did not have access to safely managed services in 2022 [2]. Nearly half of this rural population without safely managed services also lacked basic services, defined by the United Nations Joint Monitoring Programme (UN JMP) as failing at least one of the safely managed criteria while still employing an improved source with a collection time of 30 minutes or less.

Despite recent progress, the goal of universal safely managed services remains far off in many low-resource settings, including rural areas of sub-Saharan Africa. For example, only 19% of rural residents in Ghana had safely managed services in 2022 (up from 11% in 2015), while 26% lacked basic services [2]. The most recent national census (2021) has shown that access levels in some northern regions are especially poor: <50% of rural households in Northern, Savannah, and North East regions have basic access, while the Upper West and Upper East regions performed better (71-75% basic access among rural households) [3]. Economic conditions likely contribute to these poor water services, as northern Ghana tends to have higher levels of poverty compared with other parts of the country. Approximately seven out of ten residents fall below Ghana’s upper poverty line in northern regions [4].

Where gaps in safely managed services are large, such as in rural northern Ghana, a complete, single-step transition to services that meet all three safely managed criteria may be unlikely. Instead, the transition may occur more gradually over multiple steps, with intermediate systems that focus on improving one or two of the key attributes (accessibility, availability, or safety). At present, it is unclear how these three attributes align with household priorities and willingness-to-pay (WTP). Knowledge is limited regarding user satisfaction with drinking water services in rural low-resource settings, particularly concerning users’ preferences, priorities, and WTP around specific characteristics and features of those systems [5,6]. Studies in higher-income contexts have found that users are willing to pay for improvements to existing water services [7–9]. Similarly, understanding which attributes households in rural low-resource settings value most would help tailor water services aligned with users’ priorities in these contexts. Services that better reflect user preferences could improve service performance and sustainability through an increased sense of community ownership, higher WTP for tariffs, and higher levels of maintenance [6,10]. Examination of these issues may also help define user-informed intermediary targets on the pathway toward services that are fully safely managed, by identifying the service features that users see as improvements over others.

Accordingly, this research explored current satisfaction levels with existing water services, likes and dislikes about those services, and interest in service improvements among households in rural northern Ghana. We focused on answering the following research questions: (i) What aspects of water services most influence users’ satisfaction with those services, and how do these trends vary based on water system, household, or community characteristics? (ii) What water service features do people value most when considering hypothetical improvements to water systems? To explore these questions, this study employed household surveys to measure user satisfaction with existing water services and WTP for various improved service features through discrete choice experiments. We also conducted community water point observations, as well as focus groups and interviews with district officials. Broadly, we hypothesized that rural households do not value all service features equally, and trade-offs in satisfaction and valuation between service attributes could provide an evidence base for intermediary steps on the service ladder towards safely-managed access.

## 2. Methods

### 2.1. Study location

This study primarily uses cross-sectional household surveys conducted during baseline data collection for a program focused on improving water and sanitation in rural northern Ghana. This baseline effort occurred in the Upper West, Upper East, Northern, and North East regions of Ghana, with survey s taking place in two districts within each region. These districts included: Wa East and Daffiama Bussie Issa from the Upper West Region, Bawku West and Tempane from the Upper East Region, Mion and Yendi from the Northern Region, and East Mamprusi and Mamprugu Moagduri from the North East Region. In each district, we began with a list of rural communities being targeted for program activities due to poor existing water and sanitation conditions (e.g., high open defecation rates), and we randomly selected 14-15 communities to include in the survey. One district in the North East Region (East Mamprusi) contained only 14 communities targeted for intervention, while all other districts included 15 communities.

In total, we surveyed 120 communities in northern Ghana. Because communities were selected based on planned program implementation, they are not necessarily representative of the study districts or regions as a whole. Instead, they represent communities with some of the poorest levels of water and sanitation access, which district officials identified as most in need of improvements. Broadly, all selected communities were rural, and most were in relatively remote locations, although a small number of rural small towns were included. Household surveys occurred from December 2022 to February 2023, during northern Ghana’s dry season, when water availability may be most challenging.

In addition to the household surveys (described in further detail below), we also conducted focus group discussions and follow-up interviews with members of the District Interagency Coordination Committee on Sanitation (DICCS). We integrated information from these activities with our household results by creating district-level variables focused on topics such as WASH-related policies, budgets, and monitoring.

### 2.2. Household and respondent selection

Within each community, we surveyed 12-15 households, for a total of 1,748 households across the entire study area. We randomly selected households according to the following procedure: At the beginning of a new survey, the CommCare survey application (used to conduct the surveys on Android phones) assigned each enumerator a randomly-generated direction (out of eight possible cardinal directions, including north, northeast, east, etc.) and a random number of households. This number was based on the community size: In communities with less than 20 households, this number could range from 1 to 10, and in communities with 20 or more households, it could range from 1 to 25. The enumerator then walked in the assigned cardinal direction until they passed the assigned number of households, and the last household they counted was asked to participate.

After selecting a household, the enumerator aimed to target an adult female involved in the household’s financial decisions. We targeted female respondents because women in this context are often most involved in household water and sanitation provision. If such an individual was not available, the enumerator surveyed another adult household member who was at least 18 years of age and was knowledgeable about financial decisions. Prior to beginning the survey, the enumerator explained the study, confirmed the participant’s age, and obtained written informed consent.

### 2.3. Survey design

The baseline survey included numerous questions concerning water, sanitation, and hygiene. For the present study, we asked about households’ current water supply characteristics, practices, and payments, as well as levels of satisfaction with these existing services and willingness-to-pay (WTP) for hypothetical alternative services. To assess satisfaction, we asked about respondents’ overall levels of satisfaction with respect to their current water service using a four-point Likert scale (very unsatisfied, somewhat unsatisfied, satisfied, very satisfied). We also asked follow-up questions regarding which aspects of the service (accessibility, availability, reliability, safety, and affordability) they liked or disliked.

To estimate WTP for water services with various hypothetical characteristics, we used discrete choice experiments (DCE, described in detail in the following section) among a random subset of baseline survey respondents (N = 542, 31% of all baseline respondents), as this portion of the survey required a considerable amount of time to complete. In addition to the DCE implementation within the baseline survey, we also conducted 424 supplemental DCE surveys in northern Ghana focused specifically on small town residents with greater exposure to piped water services. These additional surveys enabled us to investigate whether this slightly different context would alter how individuals valued the water service features included in the hypothetical DCE scenarios. The supplemental DCE surveys followed a similar approach for random household selection with modifications to ensure that both on-premises and off-premises water users were captured. These surveys took place in May-June 2023 in the Upper West, Upper East, North East, and Savannah regions.

In addition to household surveys, enumerators also observed community water points. For each community water point, enumerators completed a separate CommCare form focused on water point infrastructure, functionality, and upkeep. Key water point observations included whether water points were functional, well-maintained, and clean at the time of data collection, and whether they were easily accessible (e.g., not obstructed by vegetation or debris).

### 2.4. Discrete choice experiment design and implementation

The DCE method enabled us to consider the impact of separate features reflecting different types of improvements in moving toward safely managed water services (Table 1). In this approach, the enumerator presented a survey respondent with a series of hypothetical choices between two scenarios. Each scenario described a combination of five service features (Table 1, SI Table S1). The two scenarios presented to the respondent are known together as a choice set (see SI Figure S1 for an example).

**Table 1.** Water service features considered in discrete choice experiments to estimate willingness-to-pay. Prices are shown in Ghanaian Cedis (GHS; 11 GHS = 1 USD in 2023 during data collection).

| Feature | Feature levels | Description and notes |
| --- | --- | --- |
| Source type and accessibility | • piped in home<br>• piped on plot<br>• public standpipe within 10 minutes<br>• public handpump within 10 minutes<br>• public handpump within 20 minutes<br>• public handpump within 30 minutes | Source type and accessibility are considered together because certain sources are more likely to have increased accessibility (e.g., in-home water is almost certainly piped); longer distances were only included for handpumps for simplicity. |
| Water quality & treatment | • chlorinated<br>• unchlorinated | For simplicity, we focused on chlorination, as many respondents were familiar with this treatment. |
| Availability | • full (water on 24 hours per day)<br>• limited (4 scheduled hours per day) | We defined “availability” as the degree to which the system needs to ration water by scheduling regular downtime; we included only two levels, defined in consultation with local experts, for simplicity. |
| Reliability | • high (95%: 1-2 service interruptions per month during scheduled time, with breakdowns addressed in 24 hours)<br>• low (70%: service interruptions at least once a week, with breakdowns lasting for more than 24 hours) | We defined “reliability” as the degree to which the system is operational when it is scheduled to provide water; we included only two levels, defined in consultation with local experts, for simplicity. |
| Price & affordability | • 2.5 GHS per m<sup>3</sup> (or 0.05 GHS per 20-L bucket)<br>• 10 GHS per m<sup>3</sup> (or 0.20 GHS per 20-L bucket)<br>• 20 GHS per m<sup>3</sup> (or 0.40 GHS per 20-L bucket)<br>• 35 GHS per m<sup>3</sup> (or 0.70 GHS per 20-L bucket) | We presented the water tariff associated with a given scenario using both scales (per m<sup>3</sup> and per 20-L bucket), so respondents could more easily compare with their current approach to water collection and payments; included tariff values were designed to encompass the range of likely WTP responses, based on survey pre-testing in the region. |

A full factorial design, that would include all possible combinations of the features, would have required 192 distinct scenarios and over 18,000 unique choice sets. Instead, we employed a Fedorov exchange algorithm [11] to estimate an optimal partial design that considered: (i) orthogonality, minimizing the possible correlation between features, and (ii) level balancing, ensuring that the levels within each feature appeared a similar number of times. We ran the algorithm multiple times to assess possible partial designs containing up to 30 scenarios, repeating the procedure up to 1,000 times at each potential sample size to ensure it arrived at the best possible outcomes. The selected design contained 24 scenarios (SI Table S1), resulting in a total of 276 unique choice sets. Finally, we removed 35 choice sets where one scenario was superior or equivalent to the other with respect to every feature (including price), as we would always expect respondents to choose the superior scenario (providing no useful information for understanding tradeoffs). This process left us with a total of 241 choice sets included in the survey.

After walking a respondent through a simple introductory example (involving a choice of two meals), the enumerator then presented a series of four choice sets, each randomly selected by the CommCare application. A set of 24 informational handouts aided the enumerator in describing the two scenarios within each choice set (examples provided in SI Figure S1). Following the explanation of a choice set, the respondent stated which scenario they would prefer. In our survey, they were also able to opt out of the choice, meaning they would not pay anything but not gain access to either scenario (in this hypothetical situation). After completing the four choice sets, the enumerator repeated the first choice set as a quality assurance check, confirming that the participant responded in the same way. Only 3% of respondents were inconsistent in choice.

In addition to the DCE choice sets, we also asked open-ended questions about WTP for two specific scenarios from our set of 24. These two scenarios represented the highest and lowest overall levels of service:

- Highest level of service (Scenario 1 in SI Table S1): What is the maximum amount that your household would be willing to pay for access to chlorinated water from a tap inside your dwelling, where the water service is available 24 hours per day and is interrupted no more than 1-2 days per month, and breakdowns never last more than 24 hours per day because someone performs regular maintenance?
- Lowest level of service (Scenario 6 in SI Table S1): What is the maximum amount that your household would be willing to pay for access to unchlorinated water from a public handpump, where it takes 30 minutes to go, collect water, and return home; the water is typically available for 4 scheduled hours per day; water service is interrupted 2 days per week and breakdowns can last more than 24 hours because maintenance only happens when needed?

WTP for the highest level of service was used as a consistency check for DCE responses. If the respondent had selected DCE scenarios with higher price points than their WTP for the highest level of service, the enumerator went back and confirmed those answers. After going back, respondents confirmed their selection of higher price points than their maximum reported WTP in 10% of choice sets. We removed these discrepancies during analysis. WTP for the lowest level of service was used as a reference level, allowing us to add on WTP increases for specific service features derived from the DCE analysis.

### 2.5. Data analysis

#### 2.5.1. Household, community, and district characteristics

We calculated descriptive statistics on key household survey topics, such as water service satisfaction, drinking water source, and respondents’ likes and dislikes about their current service. We used information about water sources and conditions to infer whether households had access to basic or safely managed service levels, according to UN Joint Monitoring Programme (JMP) criteria [1]. We also derived wealth quintiles for our study population according to the wealth index methodology used by the Demographic and Health Survey Program [12]. We created wealth quintiles specific to our survey population for easier comparisons of wealth across respondents, because 93% of our baseline respondents fell within rural Ghana’s poorest quintile.

We used a combination of aggregated household survey responses and community water point observations to develop an understanding of community-wide characteristics. We used aggregated household responses primarily to better understand the overall socioeconomic condition of the community. For example, we determined what fraction of households in a given community fell in the bottom or top wealth quintile of our entire study population, as well as what fraction had heads of household who were female, who had no formal education, or who were employed in agriculture (SI Table S2).

In addition to household and community characteristics, we also reviewed focus group discussions and interviews with district officials to identify whether district-level activities were occurring that may improve WASH conditions in communities. For example, these activities included whether the district had conducted WASH needs assessments, monitored piped systems, or implemented water governance accountability mechanisms (SI Table S2).

#### 2.5.2. Factors associated with existing water service satisfaction levels

Using these household, community, and district characteristics, we examined relationships with survey respondents’ satisfaction with their existing water services. In particular, we performed a multivariate logistic regression analysis with satisfaction as the dependent variable. For the purpose of this logistic regression, we simplified the survey’s four-level satisfaction scale to a binary indicator of whether respondents were satisfied (including “very satisfied” or “satisfied” responses from the survey) or unsatisfied (“somewhat unsatisfied” or “very unsatisfied”)

We began with a large model containing 10 household, 17 community, and 9 district-level explanatory variables, along with one additional variable representing the region (SI Table S2). The model also included adjusted standard errors to account for any community clustering not described explicitly by the community-level variables. We then iterated through successively smaller models by sequentially removing variables with the largest p-values, suggesting they had limited association with satisfaction. We also eliminated any variables that posed collinearity concerns, based on computed variance inflation factors (VIF) [13,14]. In the final model, all remaining variables had p<0.20 and VIF<3. When interpreting results, following guidance from the American Statistical Association [15], we avoided using a threshold p-value such as 0.05 to make definitive statements of statistical significance, although we still reported p-values to aid in interpretation.

#### 2.5.3. Willingness-to-pay for hypothetical water service features

We used a mixed logit model [16] to analyze WTP for various water service features based on our DCE data. This model contained all features considered in the DCE scenarios, including water source type and location, chlorination, availability, reliability, and price. Reference levels for each of these categorical features were set at their lowest (worst) values, so that all results would reflect WTP for improvements over the lowest hypothetical level of service (defined as a public, unchlorinated handpump requiring a 30-minute collection time, where water is typically available for 4 set hours per day, but unscheduled service interruptions occur 2 days per week and can last more than 24 hours).

All feature variables apart from price were formulated as normally-distributed random parameters, which means that estimated coefficients could vary across respondents, allowing for individual variations in preferences. We determined WTP values for a given improved feature (relative to the associated reference value) by calculating marginal utility, found by dividing that parameter’s random coefficient by the price coefficient estimated in the model. Given previous findings showing that WTP derived from stated methods (such as DCE) may overestimate revealed WTP, in which the participant pays real money [17], we interpreted results from our DCE model as indicating trends in how respondents prioritized different service features, rather than as accurate amounts they were willing to pay for these features. We used R Version 4.3.1 for all data analysis procedures.

### 2.6. Research ethics

The Institutional Review Board for the Council of Scientific and Industrial Research in Ghana reviewed and approved our research protocol (RPN 022/CSIR-IRB/2022). Prior to any field work, the study was introduced to District Assembly members, and field work supervisors described the work and requested permission from community leaders. We obtained written informed consent from all survey participants, and all collected data was kept confidential. We have shared initial results with local authorities in aggregated form, with identifiable information removed.

## 3. Results

### 3.1. Characteristics of household respondents

Across our study population, 77% of 1,748 total baseline survey respondents were female, while only 13% were members of female-headed households (Table 2; these results focus only on baseline respondents and do not include supplemental DCE participants; characteristics of supplemental participants are provided in SI Table S3). Female heads of household were slightly more common in the Upper West and Upper East regions. According to our computed wealth quintiles, Upper East households tended to be less wealthy than those in other regions. Across all regions, household heads were typically employed in agriculture and had completed no formal education. Local officials classified most communities as remote rural, but 23% of respondents lived in communities classified as rural small towns, which tended to be larger and located along main roads.

**Table 2.**
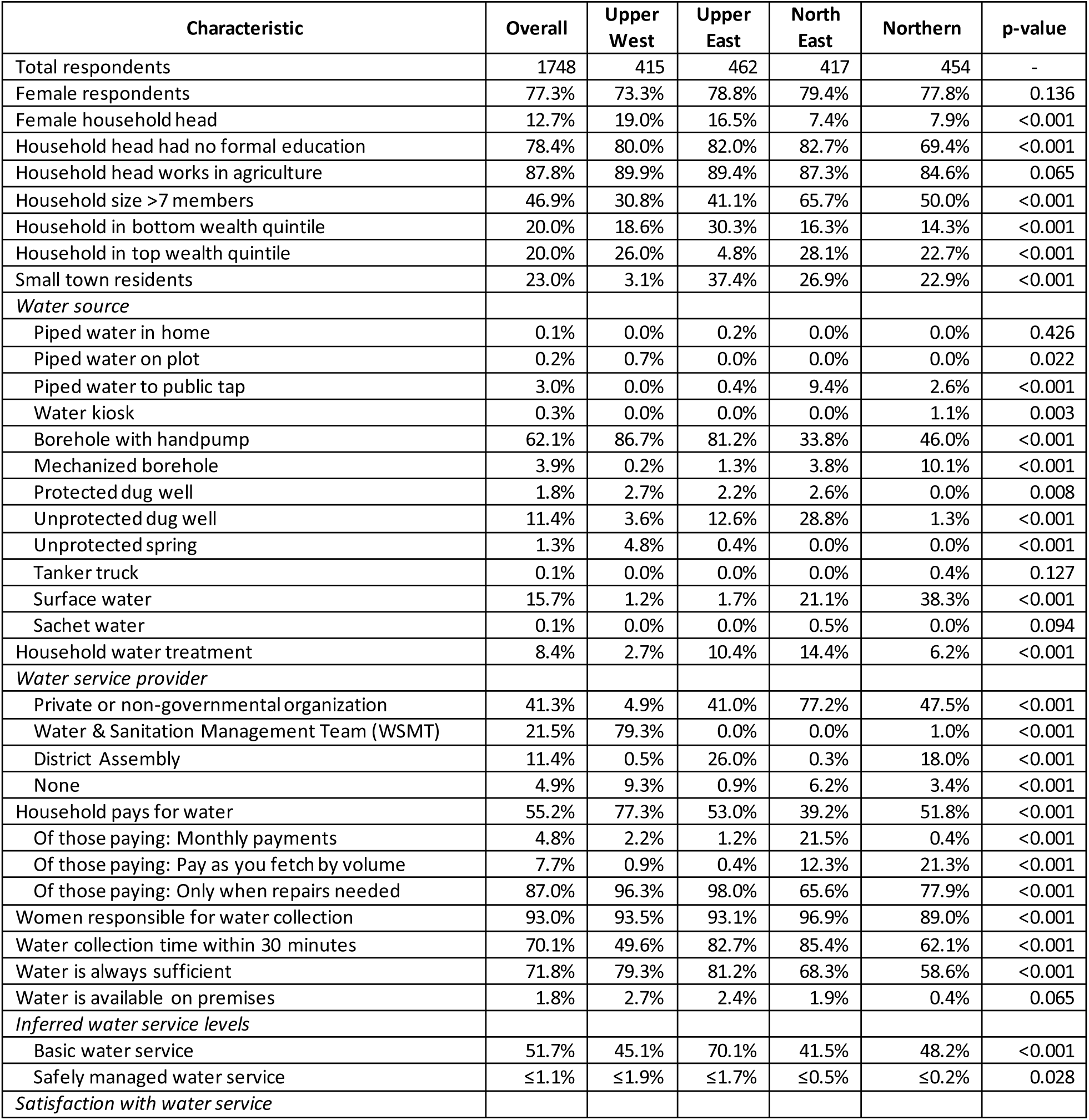
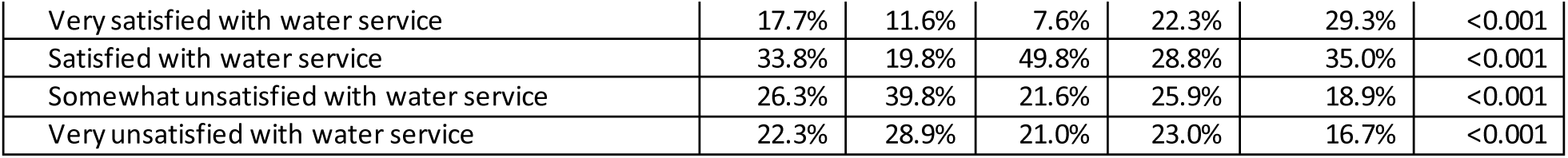
Descriptive statistics showing baseline survey respondent characteristics across the entire study population and within each of the four regions. To compare characteristics across regions, we applied chi-squared tests in R, with p-values reported in the final column. We used UN JMP definitions to define basic and safely managed water services. Because we did not have data on water source contamination, the table reports the maximum possible percentage using safely managed services.

| Characteristic | Overall | Upper West | Upper East | North East | Northern | p-value |
| --- | --- | --- | --- | --- | --- | --- |
| Total respondents | 1748 | 415 | 462 | 417 | 454 | - |
| Female respondents | 77.3% | 73.3% | 78.8% | 79.4% | 77.8% | 0.136 |
| Female household head | 12.7% | 19.0% | 16.5% | 7.4% | 7.9% | <0.001 |
| Household head had no formal education | 78.4% | 80.0% | 82.0% | 82.7% | 69.4% | <0.001 |
| Household head works in agriculture | 87.8% | 89.9% | 89.4% | 87.3% | 84.6% | 0.065 |
| Household size >7 members | 46.9% | 30.8% | 41.1% | 65.7% | 50.0% | <0.001 |
| Household in bottom wealth quintile | 20.0% | 18.6% | 30.3% | 16.3% | 14.3% | <0.001 |
| Household in top wealth quintile | 20.0% | 26.0% | 4.8% | 28.1% | 22.7% | <0.001 |
| Small town residents | 23.0% | 3.1% | 37.4% | 26.9% | 22.9% | <0.001 |
| *Water source* |  |  |  |  |  |  |
| Piped water in home | 0.1% | 0.0% | 0.2% | 0.0% | 0.0% | 0.426 |
| Piped water on plot | 0.2% | 0.7% | 0.0% | 0.0% | 0.0% | 0.022 |
| Piped water to public tap | 3.0% | 0.0% | 0.4% | 9.4% | 2.6% | <0.001 |
| Water kiosk | 0.3% | 0.0% | 0.0% | 0.0% | 1.1% | 0.003 |
| Borehole with handpump | 62.1% | 86.7% | 81.2% | 33.8% | 46.0% | <0.001 |
| Mechanized borehole | 3.9% | 0.2% | 1.3% | 3.8% | 10.1% | <0.001 |
| Protected dug well | 1.8% | 2.7% | 2.2% | 2.6% | 0.0% | 0.008 |
| Unprotected dug well | 11.4% | 3.6% | 12.6% | 28.8% | 1.3% | <0.001 |
| Unprotected spring | 1.3% | 4.8% | 0.4% | 0.0% | 0.0% | <0.001 |
| Tanker truck | 0.1% | 0.0% | 0.0% | 0.0% | 0.4% | 0.127 |
| Surface water | 15.7% | 1.2% | 1.7% | 21.1% | 38.3% | <0.001 |
| Sachet water | 0.1% | 0.0% | 0.0% | 0.5% | 0.0% | 0.094 |
| Household water treatment | 8.4% | 2.7% | 10.4% | 14.4% | 6.2% | <0.001 |
| *Water service provider* |  |  |  |  |  |  |
| Private or non-governmental organization | 41.3% | 4.9% | 41.0% | 77.2% | 47.5% | <0.001 |
| Water & Sanitation Management Team (WSMT) | 21.5% | 79.3% | 0.0% | 0.0% | 1.0% | <0.001 |
| District Assembly | 11.4% | 0.5% | 26.0% | 0.3% | 18.0% | <0.001 |
| None | 4.9% | 9.3% | 0.9% | 6.2% | 3.4% | <0.001 |
| Household pays for water | 55.2% | 77.3% | 53.0% | 39.2% | 51.8% | <0.001 |
| Of those paying: Monthly payments | 4.8% | 2.2% | 1.2% | 21.5% | 0.4% | <0.001 |
| Of those paying: Pay as you fetch by volume | 7.7% | 0.9% | 0.4% | 12.3% | 21.3% | <0.001 |
| Of those paying: Only when repairs needed | 87.0% | 96.3% | 98.0% | 65.6% | 77.9% | <0.001 |
| Women responsible for water collection | 93.0% | 93.5% | 93.1% | 96.9% | 89.0% | <0.001 |
| Water collection time within 30 minutes | 70.1% | 49.6% | 82.7% | 85.4% | 62.1% | <0.001 |
| Water is always sufficient | 71.8% | 79.3% | 81.2% | 68.3% | 58.6% | <0.001 |
| Water is available on premises | 1.8% | 2.7% | 2.4% | 1.9% | 0.4% | 0.065 |
| *Inferred water service levels* |  |  |  |  |  |  |
| Basic water service | 51.7% | 45.1% | 70.1% | 41.5% | 48.2% | <0.001 |
| Safely managed water service | ≤1.1% | ≤1.9% | ≤1.7% | ≤0.5% | ≤0.2% | 0.028 |
| *Satisfaction with water service* |  |  |  |  |  |  |
| Very satisfied with water service | 17.7% | 11.6% | 7.6% | 22.3% | 29.3% | <0.001 |
| Satisfied with water service | 33.8% | 19.8% | 49.8% | 28.8% | 35.0% | <0.001 |
| Somewhat unsatisfied with water service | 26.3% | 39.8% | 21.6% | 25.9% | 18.9% | <0.001 |
| Very unsatisfied with water service | 22.3% | 28.9% | 21.0% | 23.0% | 16.7% | <0.001 |

Most households (62%), especially those in the Upper West and Upper East regions, reported using a borehole with a handpump as their primary water source (Table 2). Surface water and unprotected dug wells, both of which are classified as unimproved sources, were the second (16%) and third (11%) most common sources, respectively. The Northern region exhibited the highest percentages of surface water use (38%), while the greatest use of unprotected dug wells occurred in the North East (29%). Overall, piped water was uncommon (<4%), with only 4 respondents having access to piped water on premises (on plot or in home). However, nearly 10% of North East respondents reported using piped water from public taps. More than half of respondents reported paying for water services, although most of these (87%) only paid when repairs were needed.

We classified approximately half of respondents (52%) as having access to at least basic water services (Table 2). Most respondents used improved sources such as boreholes, and a majority (70%) reported collection times within 30 minutes, which is also necessary for basic access. Collection times tended to be longer in the Upper West region (only 50% within 30 minutes). Across the study area, respondents who used surface water, unprotected springs, tanker trucks, or water kiosks also tended to have longer collection times (50% within 30 minutes for this group of sources, compared with 74% for others). In 93% of households, adult women were largely responsible for the task of collecting water.

We classified very few households (<2%) as having access to safely managed water services, primarily because only 2% met the safely managed criterion of accessibility on premises (Table 2). The availability criterion was less limiting, with most respondents (72%) reporting that sufficient drinking water was always available when needed. Our results do not provide direct evidence of water quality and the third safely managed criterion (free from contamination). However, only 8% of respondents reported treating their water.

Despite having water services that rarely met more than one safely managed criteria, approximately half of household respondents (51%) reported being satisfied or very satisfied with their current services, although satisfaction levels were considerably lower in the Upper West region (Table 2). In the following sections, we explore the factors that were associated with satisfaction, as well as the relative value respondents placed on these factors, through analysis of (i) existing water services and satisfaction levels, and (ii) hypothetical scenarios with varying service features. Insights gained through these analyses could provide valuable information regarding how to progress toward safely managed services along a path that is most aligned with user priorities.

### 3.2. Factors associated with existing satisfaction levels

An initial comparison of satisfaction levels and water source showed that households tended to be more satisfied when they had access to an improved water source (Figure 1). Satisfaction was low among those using surface water (26% satisfied or very satisfied), as well as unprotected dug wells (32%) and unprotected springs (0%). In contrast, satisfaction was particularly high among those with piped water access (81% satisfied or very satisfied), even when off premises (79%). Satisfaction was also high among those using boreholes (61%), especially for the 4% with access to mechanized boreholes (i.e., where the person fetching does not have to pump; 81% satisfied or very satisfied).

**Figure 1.**
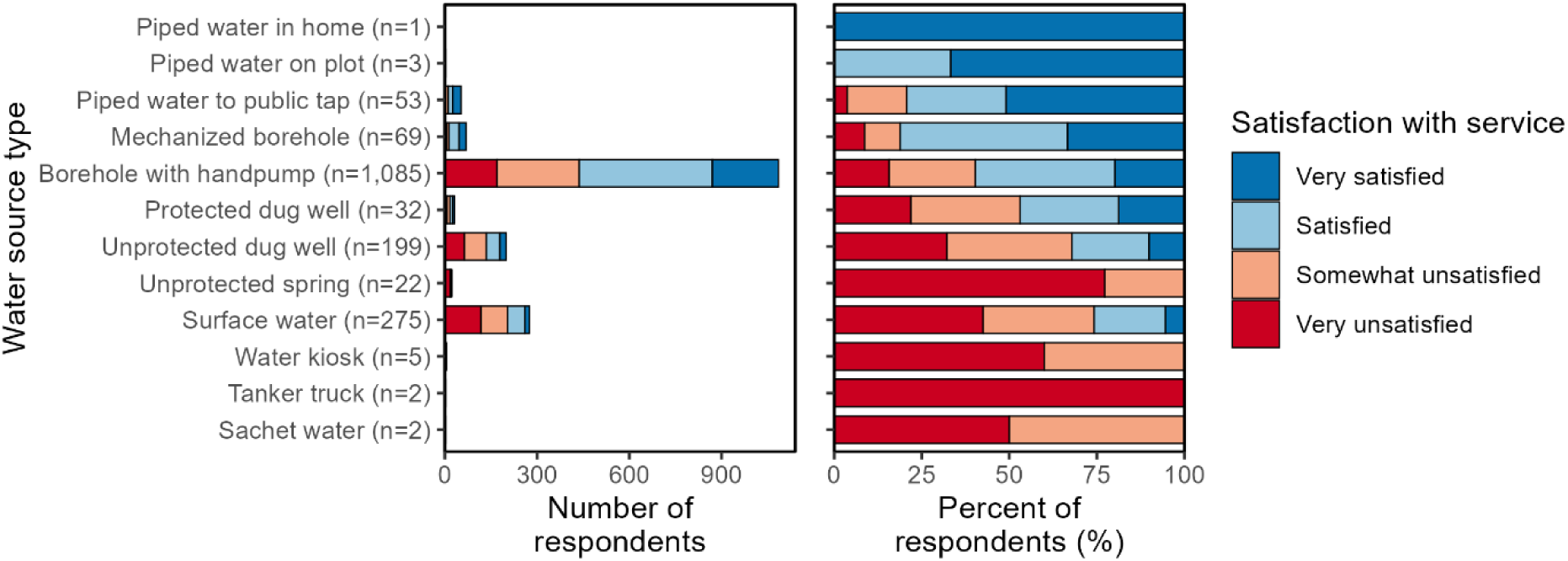
Level of satisfaction with water services for household respondents using different types of water sources for drinking. To more easily visualize the relative number of respondents using each source type as well as the satisfaction levels associated with each source, the graphs provide both the number and percentage of respondents in each category.

Multivariate logistic regression confirmed these trends when accounting for other variables (Figure 2). Unimproved water sources such as unprotected dug wells and surface water were associated with lower odds of satisfaction than boreholes with handpumps (the most common source type) and other improved sources. For example, using surface water reduced the odds of being satisfied by 94%, compared with hand-powered boreholes (odds ratio [OR]: 0.062, 95% CI: 0.033-0.116, p<0.001). Unprotected dug wells had a similar effect (OR: 0.146, 95% CI: 0.089-0.242, p<0.001), while protected dug wells were also associated with somewhat lower satisfaction than other improved sources (OR: 0.431, 95% CI: 0.207-0.899, p=0.025). Notably, we were unable to include certain source types, including piped water on premises, unprotected springs, and sachet water, in this regression model, because they were used by very few respondents.

**Figure 2.**
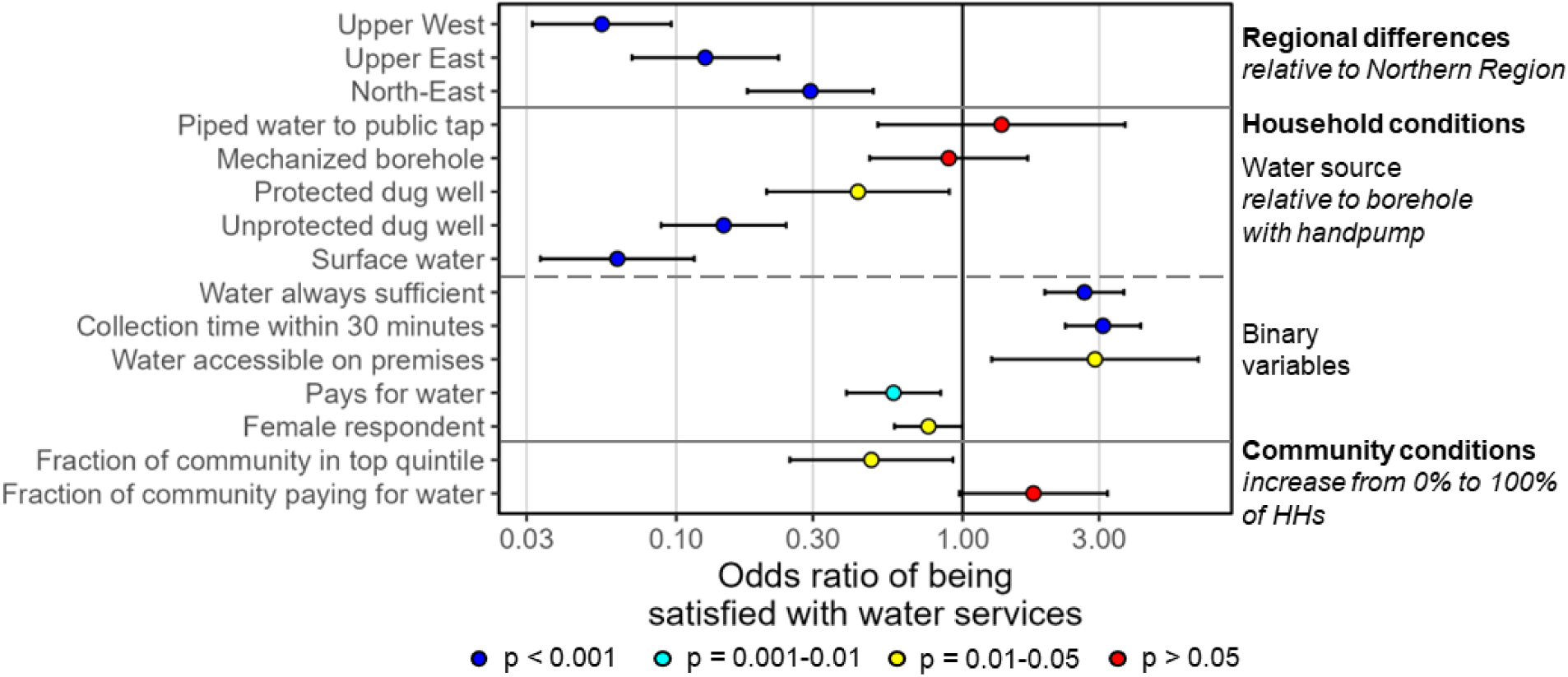
Variables associated with water service satisfaction among 1,748 households, based on logistic regression. For the purpose of this regression, the four-level Likert scale was consolidated into two levels: satisfied (including “satisfied” and “very satisfied”) and unsatisfied (including “somewhat unsatisfied” and “very unsatisfied”). All variables shown were included in the same multivariate regression model, which included adjusted standard errors to account for community clustering. Additional variables were considered in earlier models (SI Table S2), and variables with limited association with satisfaction were removed sequentially until the model only contained variables with p<0.20. Variables exhibiting high levels of collinearity (variance inflation factor >5) were also removed. Variables with odds ratios above 1 are positively associated with satisfaction, while odds ratios below 1 are negatively associated. Error bars represent 95% confidence intervals. Reference levels for categorical variables with more than two possible values (region, water source) are defined to the right of the graph. Variables are grouped by scale (region, household, community).

Beyond the type of water source, satisfaction levels were also associated with several other factors, some of which corresponded to criteria for safely managed or basic access. Satisfaction levels tended to be higher among households who met the availability criterion, reporting that they always had drinking water available in sufficient quantities (OR: 2.66, 95% CI: 1.94-3.66, p<0.001). The degree of water source accessibility also seemed to be an important consideration for our survey population. Respondents able to collect water within 30 minutes had over three times the odds of being satisfied, compared with those who were further away (OR: 3.09, 95% CI: 2.28-4.19, p<0.001). Additionally, those few with access to water on premises had higher satisfaction, although the small sample size limited our ability to precisely estimate the associated odds ratio (OR: 2.90, 95% CI: 1.26-6.66, p=0.012).

With respect to household characteristics, satisfaction levels tended to be somewhat lower among female respondents (OR: 0.760, 95% CI: 0.578-0.999, p=0.049). As women were primarily responsible for water collection in 93% of our survey households, they may be more aware of water source availability and maintenance issues than men. Poorer water services may also require more of women’s effort and time. Households that reported paying for water (in most cases for maintenance and repairs) also tended to be less satisfied (OR: 0.574, 95% CI: 0.393-0.838, p=0.004), perhaps either because they didn’t want to spend money on water or because they felt their payments were not resulting in a meaningful improvement in service quality. However, of the households paying for water, most (87%) reported only doing so when repairs were needed.

Additionally, those living in communities where a higher fraction of households paid for water also may have had somewhat higher satisfaction (OR: 1.77, 95% CI: 0.975-3.21, p=0.061). In these communities, it may be that water service quality and maintenance were noticeably better. Interestingly, though, a respondent’s satisfaction tended to be lower if they lived in a wealthier community, where a greater fraction of households fell in the top wealth quintile relative to the rest of the study population (OR: 0.480, 95% CI: 0.249-0.926, p=0.028). Since most water sources in these areas are community water points serving multiple households, perhaps households living in wealthier communities felt that their community should be able to afford a better water system. Additionally, as small towns tend to be wealthier than more remote communities (p<0.001), it is also possible that small town residents aspired to more “urban” water systems, such as a piped supply distributed to on-premises connections. The small town variable itself was not included in the final model, because it was highly correlated with other variables such as community wealth (p<0.001), and it was not associated with satisfaction when controlling for these other variables.

At a regional level, satisfaction levels in the Upper West and Upper East regions, and to a lesser extent in the North-East region, were considerably lower than in the Northern region. Notably, these trends were contrary to results from the 2021 Census, which reported that the Northern region was worse than the Upper West and Upper East in terms of basic service access and collection time. As noted previously, our study population was not a representative sample of these regions. Communities were selected specifically for poor existing water and sanitation conditions. Since, according to the Census, the Upper West and Upper East have higher water access levels overall, the communities we sampled from those regions were likely worse off relative to their neighbors, potentially leading them to feel less satisfied with their water services. In contrast, selected communities in the Northern region may have been more similar to their region as a whole. Our results provided some corroborating evidence for this explanation, as access to basic services in the Upper West (45%) was far below the Census value for rural residents in that region (71%). In the Northern region, our results (48%) were similar to the Census data (46%).

Finally, we were unable to include local types of water service provider directly in our regression model, because this variable was highly correlated with region. However, we performed a separate regression focused solely on this variable’s association with satisfaction (SI Figure S2). Relative to private and non-governmental safe water enterprises (the most common provider type among our respondents), community Water and Sanitation Management Teams (WSMTs) were associated with lower satisfaction (OR: 0.346, 95% CI: 0.238-0.484, p<0.001). In Ghana, WSMTs are composed of community members and often have direct responsibility for maintaining rural community water systems, with oversight from District Assemblies [5]. Nearly all households served by WSMTs (99%) were located in the Upper West region, likely contributing to the lower satisfaction levels observed there. Water systems directly managed by District Assemblies had higher satisfaction levels (OR: 1.95, 95% CI: 1.17-3.23, p=0.010). District-managed systems were located primarily in the Upper East (68%), although 30% were in the Northern region. Finally, and perhaps unsurprisingly, satisfaction levels were lower when water systems had no defined manager (OR: 0.331, 95% CI: 0.141-0.774, p=0.011). The Upper West region contained approximately half of these systems.

### 3.3. Preferences concerning existing aspects of water service quality

As a complement to the regression analysis, we also analyzed how satisfaction levels related to direct statements made by respondents regarding what they liked and disliked about their existing water services. Immediately after respondents stated their satisfaction level in the survey, they were asked to select the aspects of their water service they liked, and then the aspects they disliked. The aspects they could select included accessibility, affordability, availability, safety, and reliability, and they had the option of selecting no or multiple items.

Accessibility and safety appeared to be most closely related to overall satisfaction levels. When people were unsatisfied, we saw frequent mention of disliking these service aspects, while satisfied respondents often reported liking these aspects about their water service (Figure 3). For example, 51% of satisfied respondents reported liking their water service’s accessibility, and 68% liked its safety. Conversely, 60% of unsatisfied respondents disliked accessibility characteristics of their water service, while 56% disliked its perceived safety (or lack thereof). In other words, these aspects seemed to be most important to people and may have had the most influence over whether or not they felt satisfied with their current service. It is important to note that respondents were focused on accessibility in terms of how easily the source could be reached and the absence of obstructions, rather than whether or not it was located on premises.

**Figure 3.**
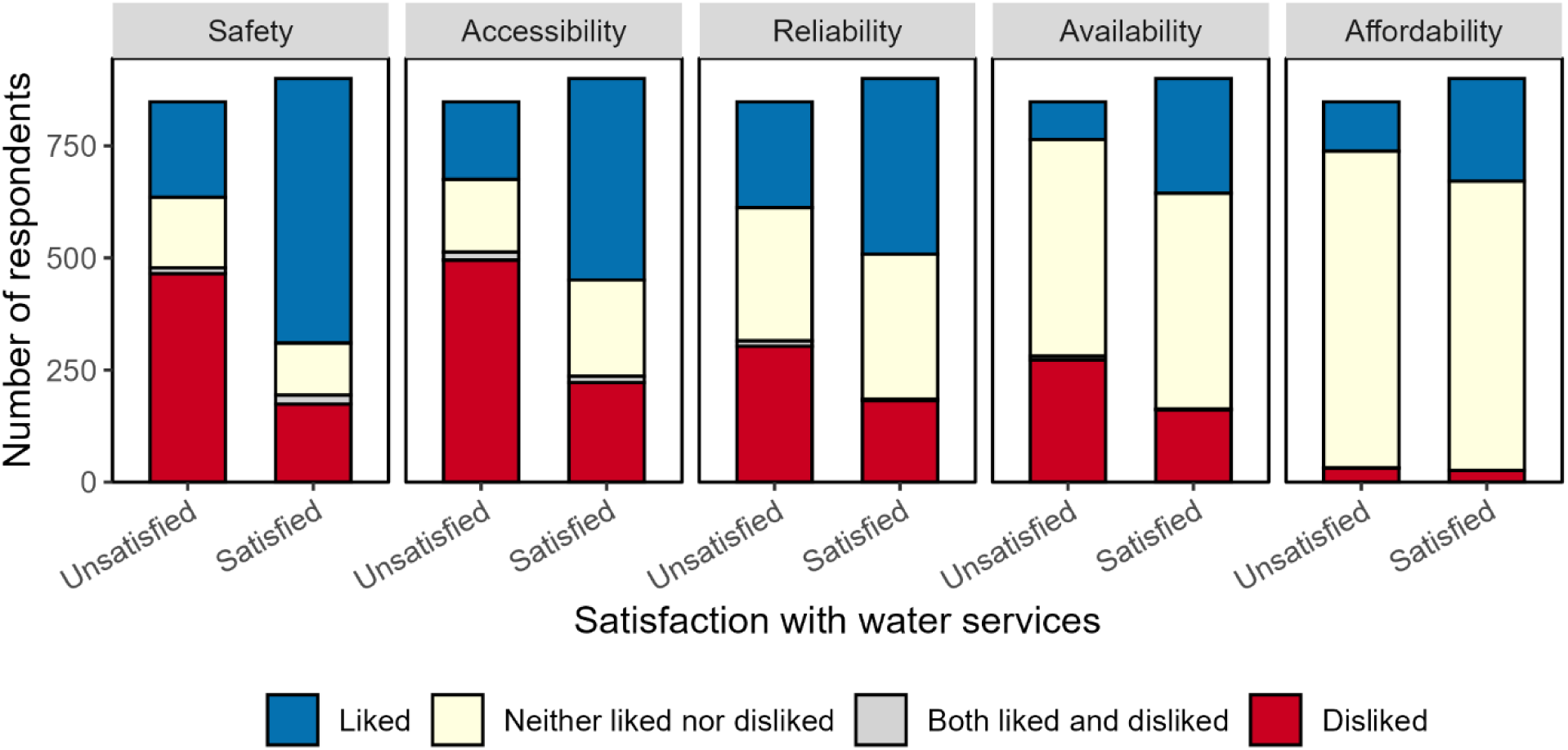
Relationships between household satisfaction with water services and the aspects of water service quality that households mentioned liking or disliking about their current service. Each plot shows how many households within each satisfaction level reported liking or disliking a specific aspect about their water service. For simplicity, we consolidated satisfaction into two levels: satisfied (including “satisfied” and “very satisfied”) and unsatisfied (including “somewhat unsatisfied” and “very unsatisfied”).

We saw similar trends with availability and reliability (Figure 3). However, the large proportions of respondents who did not mention liking or disliking these aspects suggested that they were less critical, having a smaller effect on satisfaction levels. The results also suggested few affordability concerns, with very few people mentioning disliking their water service’s level of affordability. This may relate to the fact that many respondents only made payments when the system needed maintenance or repair, but it also might suggest that respondents may be willing and able to pay for higher-quality services.

### 3.4. Preferences and willingness-to-pay for hypothetical water services

Beyond considering existing conditions, we also used DCE methods to explore respondents’ preferences around hypothetical changes to water services. In this section, we focus first on DCE results from our baseline survey (N=542), and then we discuss results from additional DCE surveys in rural small towns with greater levels of piped access (N=424). In many cases, we discuss our results in relation to the reference scenario (consisting of a public, unchlorinated handpump requiring a 30-minute collection time, where water is typically available for 4 set hours per day, but unscheduled service interruptions occur 2 days per week and can last more than 24 hours), which represented the lowest level of service among our 24 DCE scenarios. The features of our reference scenario also aligned well with typical conditions among our baseline survey population, such as the common use of handpumps with collection times of approximately 30 minutes. Mean stated WTP for this reference scenario was 8 GHS/m^3^ among baseline participants.

Broadly, despite high levels of poverty in our baseline study area, model results suggested that people would be willing to pay more for improved service quality (Figure 4). However, it is important to remember that the results presented here reflect participants’ stated preferences when no real money was involved, and previous research has found that these types of methods can overestimate respondents’ revealed WTP, generally considered to be more reflective of reality [17]. While we report estimated WTP values below, we primarily use these to inform our understanding of how respondents prioritize different service features relative to one another, rather than as recommendations for tariff increases associated with such improvements.

**Figure 4.**
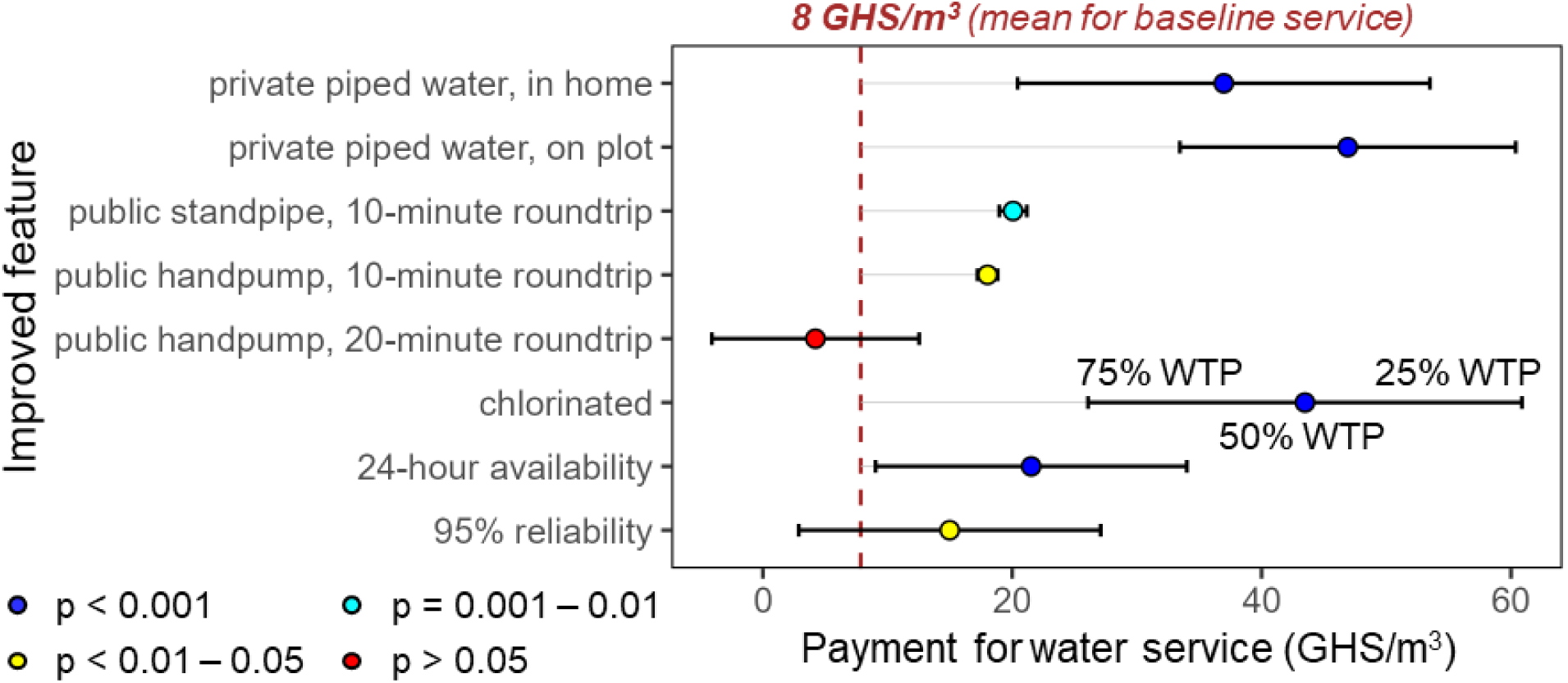
Estimated willingness-to-pay for service improvements relative to a baseline level of service (defined as access to an unchlorinated public handpump that is a 30-minute roundtrip from the household, with water available 4 hours per day and breakdowns 30% of the time), based on discrete choice experiments among baseline survey participants (N=542). The dashed red line shows mean WTP for the reference scenario, as stated by respondents in response to an open-ended question. Points represent estimates of mean WTP when an improved feature is added, and error bars represent the first and third quartiles of the modeled WTP distribution (modeled as a normal distribution based on individual preferences). Accordingly, results where error bars cross the baseline mean (e.g., 95% reliability) suggest that at least 25% of respondents may not be willing to pay more for these features, even if a majority do value them more highly than the baseline level. Note that error bars for the public handpump are not visible because their values are extremely close to the point estimate.

The most highly valued features appeared to focus on accessibility and safety, consistent with our findings in the previous section. The largest increases in WTP over the reference scenario were associated with access to private piped water connections and the addition of chlorination (mean WTP increases: 29-39 GHS/m^3^, or 3.6-4.9 times higher than the reference scenario; p<0.001). According to the model, on-plot piped water was valued a bit more highly than in-home connections (Figure 4). One possible explanation for this unexpected result is that respondents may have been more accustomed to seeing on-plot connections in other communities. Additionally, in rural northern Ghana, the prevalence of compounds shared by multiple (often related) households, where activities such as food preparation and cooking commonly occur outside the main house, likely aligns well with on-plot services. The high preference for chlorination may reflect a considerable level of concern for the quality and safety of water. While we did not directly assess the quality of existing water services, safety arose as one of our respondents’ most frequent likes or dislikes about those services. Notably, our mixed logit model showed that variations based on individual preferences were substantial for all three of these features (as shown by error bars in Figure 4, representing the spread of estimated individual WTP values). For example, while the mean WTP increase for chlorination was 36 GHS/m^3^ (4.5 times higher than the reference scenario), we estimated that 75% of respondents would be willing to pay only 18 GHS/m^3^ more than they would for unchlorinated water (2.3 times higher than the reference scenario).

Public standpipes (p=0.007) and handpumps within 10 minutes of people’s homes (p=0.013) were less valued than private piped connections but were still seen as improvements over reference handpumps that were 30 minutes away, with mean WTP increases of 10-12 GHS/m^3^ (1.3-1.5 times higher than the reference scenario; Figure 4). Notably, these two source types also exhibited much less variation in individual preferences. The random parameters associated with individual variations in these coefficient values were not statistically meaningful (p>0.90), meaning that most respondents’ estimated WTP was very close to the mean. In contrast, respondents did not exhibit increased WTP for handpumps when collection times were 20 minutes rather than 30 minutes (p=0.32).

Respondents also valued improvements to availability (mean WTP increase: 14 GHS/m^3^, or 1.8 times higher than the reference scenario; p<0.001) and reliability (mean WTP increase: 7 GHS/m^3^, or 88% higher than the reference scenario; p=0.011), although these features did not seem to be as critical as those related to accessibility and safety. WTP for availability and reliability features also varied considerably based on individual preferences (p=0.007-0.009). For example, approximately 35% of respondents would not be willing to pay anything more to improve service reliability over the reference scenario, according to the WTP distribution estimated by our model (Figure 4). However, existing water service and respondent characteristics had minimal impact on individual variations in WTP for certain features, suggesting the importance of personal preference (SI Figure S3).

In addition to these results from the baseline survey, we also conducted 424 supplemental DCE surveys in Northern Ghana focused specifically on small town residents with greater exposure to piped water services (see SI Table S3 for a comparison of these additional respondents with those from the baseline survey). The additional surveys applied the same approach used during the baseline, and they enabled us to investigate whether this slightly different context would alter how individuals valued the water service features included in the DCE scenarios.

Interestingly, results followed trends very similar to what we saw in the baseline data (SI Figure S4). Private piped water and chlorination remained the most highly valued features, while public standpipes and handpumps within 10 minutes, as well as improved availability and reliability, were less valued but still seen as worthwhile enhancements over the reference scenario. We did observe some minor differences from these additional small town results. Private piped connections (especially on-plot) were valued more highly than chlorination, and public standpipes were valued more highly than handpumps. These differences may suggest that people’s appreciation of piped water becomes even more pronounced once they have seen or experienced it more directly. In most cases, WTP values for these improved services tended to be larger than those estimated from the original baseline surveys. Except for improved availability and public handpumps within 20 minutes, where mean WTP did not increase, mean WTP was 17-86% larger than in the original surveys. These differences may be associated with the fact that the supplemental small town respondents tended to be wealthier than our original respondents (SI Table S3).

Finally, across both sets of DCE results, participants selected the “opt out” response 17% of the time (out of 3,860 total choice sets asked to 965 participants during baseline and supplemental surveys). Opting out means that the respondent did not want to choose either of the two scenarios presented in a choice set. Such a response may suggest that (i) the respondent is happy with their current water services or (ii) they were not willing to pay the prices associated with the available scenarios in the given choice set. Participants opted out most frequently (40% of the time) when both scenarios in the choice set were priced at the highest level (35 GHS/m^3^), which supports the second explanation. This result may suggest caution against setting tariff rates too high, even for the highest-quality services, as a substantial portion of our study population might be unwilling or unable to pay. We observed similar trends when considering each set of surveys individually (e.g., baseline respondents opted out in 16% of choice sets, while supplemental participants opted out 20% of the time).

## 4. Discussion

Overall, most of our study population relied on improved sources (primarily boreholes with handpumps) for their drinking water, with respondents typically reporting that water from these sources was available in sufficient quantities and collection times were within 30 minutes. Respondents who reported these positive characteristics tended to be among those who were satisfied or very satisfied with their existing water services. Additionally, users’ reported likes and dislikes, as well as their responses to hypothetical DCE scenarios, indicated that water quality (achieved through treatment approaches such as chlorination) and on-premises accessibility were key priorities, even though on-premises piped connections were rare among our study population. These findings suggest that there may be considerable interest and WTP for improved services that incorporate these features. However, the sustainability of such services, and continued satisfaction with them, would likely depend on the application of proactive payment schemes that go beyond asking users for funds only when system repairs are needed. The following sections further explore the implications of these findings.

### 4.1. Satisfaction levels and user preferences across studies and settings

Broadly, we found that approximately half of our respondents were satisfied with their existing water services, which may be somewhat higher than anticipated, since we targeted communities with relatively poor water and sanitation conditions. Based on responses concerning both existing services and hypothetical improvements, respondents seemed to value factors related to accessibility and safety most highly, although availability and reliability also played a role. A similar study conducted among communities in two districts of the Northern and Upper West regions in Ghana found that users were often dissatisfied with long collection times [5], suggesting similar accessibility concerns. In our case, while most respondents reported collection times below 30 minutes, DCE results suggest that users may still appreciate even shorter collection times (e.g., 10 minutes) if on-premises connections are not available.

With regard to water quality and safety, a study focused on urban water services in Chile found perceived water quality to have the largest impact on overall satisfaction, based on structural equation modeling [18]. Similarly, residents in the Municipality of New Propontida, Greece also exhibited high levels of WTP for improvements to drinking water quality, and they were also willing to pay for increased reliability (reducing service interruptions), though at a lower level than for improved quality [8]. Meanwhile, households in rural Ethiopia with access to improved water sources were more satisfied with the quality of water provided and, to a lesser extent, its availability, emphasizing that the source of water contributes to users’ perceptions of its quality [19]. These findings aligned with our result that respondents’ perceptions of water quality were highly associated with their satisfaction with existing services, with 68% of our satisfied respondents reporting quality/safety as one of the things they liked about their service (Figure 3). They also agreed with the high WTP for chlorination estimated from our hypothetical DCE scenarios.

Although our study and several of those discussed above suggest availability and reliability may be lower priorities for water users, other studies have noted their importance. For example, users of water points with increased reliability (fewer service interruptions) in central Ethiopia tended to have higher satisfaction [6]. That study also highlighted several factors related to accessibility, such as water point location, distance from the home, queuing time, and whether a guard was present. In another study focused on urban towns in southern Ethiopia, reliability had a particularly large influence on satisfaction, while accessibility was less important. A possible explanation may be that piped services were more common in this urban setting, making accessibility less of a concern. In a higher-income context, residents of Canberra, Australia were willing to increase their water bill to improve reliability through reductions in the frequency and duration of service interruptions [7]. Concerning availability, residents of urban informal settlements in Accra, Ghana exhibited higher WTP for water services that were available for 12 hours per day, rather than six hours per day [20]. However, these residents’ WTP did not increase further for 24 hours per day of service, suggesting that full availability may not be a high priority for some members of low-resource communities. In our study, while issues of reliability and availability may have been less critical for users, WTP results still suggested that they were valued. These previous studies confirmed that reliability should remain an important consideration when making service improvements.

Our results also suggested that female respondents were slightly less satisfied with their water services than men. A study focused on gender disparities concerning water-related knowledge and perceptions in rural southwestern Ghana found that men and women had distinct experiences and perceptions regarding drinking water, which could influence differences in satisfaction. Women encountered daily accessibility challenges in their role as primary water source selectors and more commonly discussed concerns with water quality and safety [21]. However, it is notable that some of these concerns revolved around the taste and odor of chlorinated water, whereas we found high interest in chlorination. Ensuring that water treatment efforts do not create new issues is an important consideration, especially if negative perceptions around chlorination exist. Additionally, women and men both discussed issues around a lack of communication from WSMTs managing water points, again suggesting that this may have been a common issue among our respondents as well.

With regard to water service providers, Kumasi & Agbemor found that users were largely satisfied with services provided by WSMTs [5], whereas we found high levels of dissatisfaction when WSMTs provided water services. Among those users in the previous study who were dissatisfied with WSMTs, their leading causes for dissatisfaction involved a lack of communication and poor maintenance [5]. These items may provide insight into why our respondents served by WSMTs often tended to be dissatisfied. Another study conducted in urban towns of Southern Ethiopia also found that service providers performed especially poorly with respect to communication with customers [22]. In an informal settlement within Accra, Ghana, residents reported a preference for water services managed by community-based committees or non-governmental organizations, likely due to low levels of public trust in the local water utility and metropolitan assembly, which manage existing water and sanitation services [23]. Meanwhile, households in rural Vietnam did not exhibit a clear preference for water services managed by the public or the private sector [24]. Taken together, these results suggest that specific context and local trust in existing institutions may matter more to user satisfaction than the general type of service provider (e.g., private, government, community-based) [23].

Finally, we found some potentially contradictory results regarding payments for water services. Those paying for water were less likely to be satisfied with their existing service, while satisfaction tended to be higher when a larger fraction of households in a community were making payments. Additionally, the affordability of existing services did not appear to be a major concern, and respondents seemed to be willing to pay more for better water service quality. This last finding runs somewhat contrary to a previous study in northern Ghana, which found resistance to paying for water because it is a basic need [5]. Although, that study also found that people were satisfied with service affordability, likely because most of them only paid when the system needed repairs, rather than paying directly for the water they collected. Our survey results suggested similar arrangements in many communities. This type of reactive maintenance payment scheme, as well as schemes based on fixed monthly or annual fees independent of water consumption, have been found to be less effective in keeping systems operational [5,25]. In contrast, payment schemes more focused on preventative rather than reactive maintenance would likely be important in maintaining acceptable levels of availability and reliability. Ensuring that users see their payments as affordable and valuable would likely depend on whether the payment system is equitable for community members with potentially different income levels, and whether users perceive that the money is being used effectively to maintain services.

### 4.2. Considering an additional rung on the JMP water service ladder: “Proximate access”

Based on our results from this context and the challenges of achieving full safely managed services in rural low-resource settings, we feel that it may be worthwhile to consider the addition of an intermediate access level that would fall between “basic” and “safely managed” on the current JMP water service ladder. Below, we evaluate possible definitions for this intermediate level, which we term “proximate access” as a preliminary suggestion.

A definition for this intermediate level could begin with the “basic” definition (an improved source with a collection time of 30 minutes or less), layering on certain “safely managed” criteria or making the “basic” definition more stringent to represent partial progress from basic to safely managed services. Using the data we have available concerning our baseline survey population, we were able to consider the impact of criteria related to accessibility (on-premises connections or shorter collection times) and availability (sufficient water is available when needed). Because water quality information was not available, we could not directly evaluate the quality criterion (“free from contamination”). However, we suggest that including this water quality criterion in the “proximate access” definition may be valuable, given that quality was a high priority for our respondents concerning both existing and hypothetical water services. For the remaining criteria, we evaluated how various possible definitions of “proximate access” would affect existing water service levels among our study population (Figure 5).

**Figure 5.**
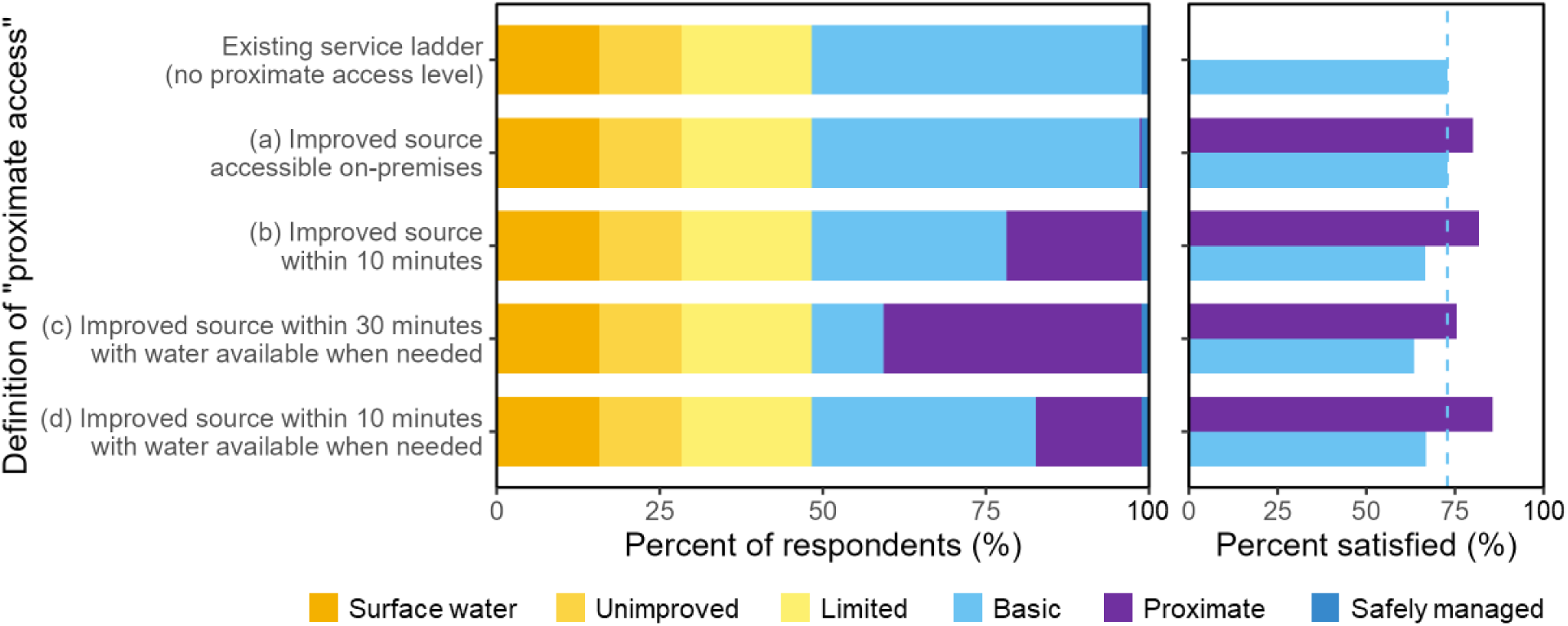
Water service ladder showing existing access levels among the baseline survey population (left), as well as satisfaction levels among those with basic and proximate access (right), using various possible definitions for the proposed proximate access level. The satisfaction plot on the right focuses on basic and proximate access only because those with proximate access always represent a subset of respondents who were classified as having basic access under the existing service ladder. The vertical dashed line indicates satisfaction among those with basic access under the existing service ladder, to provide a point of comparison. The “safely managed” category represents the maximum possible percentage of the population at this level, because our dataset did not include information on water quality.

Accessibility was highly valued by our respondents, but the small number of on-premises connections in our baseline study population confirmed that on-premises access can be challenging to achieve in remote rural settings. In conjunction, our DCE results suggested that respondents valued accessibility at three distinct levels. Improved sources within 10 minutes were valued more highly than those 20 or 30 minutes away, while on-premises connections were valued to a higher degree. This trend suggests to us a potential gap between accessibility criteria for basic access (which requires a collection time within 30 minutes) and safely managed access (which is strictly on-premises) Accordingly, we considered definitions of “proximate access” denoting access to an improved source that is (a) accessible on premises, or (b) within 10 minutes (but not on premises). Among our study population, the on-premises access criterion (a) was extremely restrictive, with <1% falling in the proximate access category (Figure 5). In contrast, the 10-minute collection time criterion (b) classified 21% of respondents as having proximate access, representing nearly half of those with basic access under the existing service ladder. In this study’s rural setting, this definition (b) may represent a substantial improvement toward safely managed services over basic services in terms of accessibility, while still not yet reaching the gold standard in full.

Respondents also valued availability and reliability, though to a lesser extent than accessibility and quality. Both availability and reliability play a role in the safely managed criterion that water is “available when needed,” so we also considered a third “proximate access” definition denoting (c) access to an improved source where water is available when needed. In our study context, this condition was frequently satisfied among our respondents, especially for those with at least basic access (79% of those with at least basic access reported sufficient water always being available, compared with 64% among those lacking basic access; p<0.001). Accordingly, this definition (c) allowed most respondents with basic access under the existing service ladder to advance to the proximate category (Figure 5). At least in our context, this definition may be too lenient to produce a meaningful difference and improvement over basic access.

Given these results, we considered combining the availability and 10-minute collection time criteria into a final possible definition (d). In this case, we classified 16% of our study population as having proximate access. Notably, this definition also resulted in the highest satisfaction level among the subset of respondents classified as having proximate access (Figure 5). Therefore, at least in the current context, we feel this definition (potentially with the added inclusion of the water quality criterion) would represent a meaningful service improvement over basic access that is feasible and valued by users.

To summarize, based on the characteristics of our study population in rural northern Ghana, we would recommend that a “proximate access” category could be defined as: access to an improved water source with a collection time of 10 minutes or less, where water is available when needed (and possibly free from contamination). Especially for rural low-resource contexts where on-premises access is a challenge, this proximate category could represent substantial progress most of the way toward safely managed services. Its inclusion in the overall service ladder could help increase motivation among local officials to continue making intermediary progress after basic access has been achieved, thereby making eventual transitions to fully safely managed services easier. However, in other, perhaps more urban settings, it is possible that the proximate category could discourage investments toward reaching safely managed services. Policy-makers and implementers would need to be careful not to see proximate access as “good enough” and deprioritize further progress.

### 4.3. Study limitations

When considering the implications of our study findings, several limitations are important to note. First, our survey sample was dependent on baseline data collection for a local water and sanitation program, which focused on locations with the greatest water and sanitation needs. Accordingly, our study population does not provide a representative picture of these regions in Northern Ghana, but it does offer a window into conditions within critical settings where considerable progress toward safely managed services is needed. We also did not measure the quality of drinking water available to survey participants, which limited consideration of the “free from contamination” criterion for safely managed services. Relatedly, a study population characterized by a greater balance between different existing water source types and accessibility levels would have helped in characterizing trends related to these factors. However, our additional DCE surveys, conducted in small towns with greater exposure to on-premises piped connections, did suggest that our baseline results may be relevant across a broader set of contexts, particularly among somewhat wealthier populations with greater existing access to on-premises piped water in northern Ghana. Potentially, greater exposure to on-premises piped services may further increase the desire for this feature.

Additionally, multiple factors limited our quantitative consideration of district-level activities, such as the implementation of water governance accountability mechanisms and the promotion of participatory decision-making. First, we were unable to include some district indicators in our regression analysis due to collinearity with the region variable, so it is possible that the regional differences we observed may partially reflect distinct district-level practices. Second, certain activities, such as documenting service providers, making annual investment plans, and improving budget systems, were only practiced by one or two districts, which limited the ability of a regression analysis to capture any effects. Finally, our interviews with district officials suggested that resource challenges surrounded regular implementation of these activities, meaning that we may not have observed an effect even for activities we did include in the regression analysis. Therefore, while our regression model did not identify any district-level variables that were strongly associated with satisfaction levels, we do not believe this finding should discourage districts from pursuing such efforts. Even when local governments are not directly providing services, they still have important roles to play in terms of management, monitoring, and oversight.

Finally, our DCE analysis relied on hypothetical scenarios and did not ask participants to pay real money. Accordingly, our results reflect respondents’ stated WTP, which may overestimate revealed WTP (i.e., how much a respondent would be willing to pay when making a real purchase). In particular, a previous study focused on WTP for sanitation in northern Ghana found that respondents’ revealed WTP averaged 53-77% of stated WTP, depending on factors such as season and whether respondents were residents of small towns [17]. In the current study, while the trends ranking the relative value of different attributes revealed through the DCE model provide valuable insight, we feel that specific WTP values from the model should be treated with caution. For example, while they might provide motivation for setting or revising tariff rates for improved services, other information should be incorporated when deciding on the precise value of those rates.

## 5. Conclusion

This study explored satisfaction with existing water services, as well as preferences and willingness-to-pay for hypothetical service improvements, among rural households and communities across four regions in northern Ghana. We found that existing satisfaction levels were highly associated with several key JMP criteria for basic and safely managed water services, including the use of improved sources (especially boreholes and piped connections) where water is always available in sufficient quantities and collection times are within 30 minutes. Criteria concerning accessibility and water quality seemed to be particularly high priorities in our study area, with respondents exhibiting high WTP for on-premises piped connections and chlorination. Meanwhile, WTP levels for improved service reliability and availability, as well as for shorter collection times (10 vs. 30 minutes) when on-premises access was not available, were lower but still meaningful. Overall, these findings supported our hypothesis that household value distinct aspects of water services to differing degrees, with especially high satisfaction and preference associated with higher-quality infrastructure such as on-premises piped connections and chlorination.

Taken together, these findings led us to consider the possibility of an intermediate JMP service level between “basic” and “safely managed”, which we have termed “proximate access”, tentatively defined as access to an improved source where collection times are within 10 minutes, water is always available, and (possibly) water is free from contamination. This proposed category may provide useful insight into service improvements that are feasible and valued in rural, low-resource settings, potentially representing an important intermediate step toward safely managed services. To promote sustainability and reliability of these services, proactive payment schemes (e.g., “pay as you fetch”) combined with effective use of funds to maintain the system and high levels of collective action (with many households contributing in a community) can help to keep services functioning and support high satisfaction levels.

## Data Availability

All data produced in the present study are available upon reasonable request to the authors.

## Acknowledgments

We thank our enumerators for their hard work during data collection, and all households who participated in our surveys. This manuscript builds on research initiated under the Enhancing Water, Sanitation, and Hygiene (EnWASH) program, which was supported by a cooperative agreement between the United States Agency for International Development (USAID) and Global Communities. We are grateful for the insight, input, and support of the EnWASH Program consortium member organizations, including Global Communities, the program lead. We also thank Water4 for their feedback and support related to the design of our willingness-to-pay surveys. The authors alone are responsible for the views expressed in this publication and they do not necessarily represent the decisions or policies of USAID or the United States Government.

