## Supplemental Information for "Satisfaction and willingness-to-pay for water services in rural northern Ghana: Do we need intermediary steps toward safely managed services?"

### Satisfaction and willingness-to-pay for water services in rural Northern Ghana: Should we expand the ladder toward safely managed services?

John Trimmer,[1,2] Joyce Kisiangani,[3] Bessy Ewoenam Odame-Boafo,[3] Lisa Appavou,[1] Chloé Poulin,[1] Dominic Osei,[4] Caroline Delaire,[1] Valerie Bauza[5*]

[1] The Aquaya Institute, Nairobi, Kenya
[2] Department of Civil & Environmental Engineering, Syracuse University, Syracuse, NY 13244, USA
[3] The Aquaya Institute, Accra, Ghana
[4] Global Communities, Accra, Ghana
[5] The Aquaya Institute, San Anselmo, California, United States of America

#### Table of Contents (11 total pages)

Supplementary Tables
SI Table S1. Scenarios used for discrete choice experiments. ........ S2
SI Table S2. Explanatory variables considered in the satisfaction regression analysis ........ S3
SI Table S3. Survey respondent characteristics from baseline and additional DCE surveys ........ S6

Supplementary Figures
SI Figure S1. Examples of informational handouts used to explain DCE scenarios ........ S7
SI Figure S2. Associations between household satisfaction and water service provider ........ S8
SI Figure S3. Estimated willingness-to-pay for water service improvements among subsets of the baseline survey population ........ S9
SI Figure S4. Estimated willingness-to-pay for water service improvements among supplemental survey respondents from small towns ........ S11

**Supplementary Information**

**Table S1.** Scenarios used for discrete choice experiments, selected using a Fedorov exchange algorithm. See Table 1 for more detail on features and levels. Prices are shown in Ghanaian Cedis (GHS; 11 GHS = 1 USD in 2023 during data collection).

| Scenario | Access type | Quality | Availability | Reliability | Price point |
|---|---|---|---|---|---|
| 1 (best) | Private in home | Chlorinated | 24 hours per day | High reliability | 35 GHS/m$^3$ or 0.70 GHS per 20-L bucket |
| 2 | Public piped, 10 min | Chlorinated | 24 hours per day | High reliability | 35 GHS/m$^3$ or 0.70 GHS per 20-L bucket |
| 3 | Public handpump, 10 min | Chlorinated | 4 hours per day | High reliability | 35 GHS/m$^3$ or 0.70 GHS per 20-L bucket |
| 4 | Public handpump, 20 min | Unchlorinated | 24 hours per day | Limited reliability | 35 GHS/m$^3$ or 0.70 GHS per 20-L bucket |
| 5 | Private on plot | Unchlorinated | 4 hours per day | Limited reliability | 35 GHS/m$^3$ or 0.70 GHS per 20-L bucket |
| 6 (worst) | Public handpump, 30 min | Unchlorinated | 4 hours per day | Limited reliability | 35 GHS/m$^3$ or 0.70 GHS per 20-L bucket |
| 7 | Public handpump, 20 min | Unchlorinated | 24 hours per day | High reliability | 20 GHS/m$^3$ or 0.40 GHS per 20-L bucket |
| 8 | Private on plot | Chlorinated | 4 hours per day | High reliability | 20 GHS/m$^3$ or 0.40 GHS per 20-L bucket |
| 9 | Public handpump, 30 min | Unchlorinated | 4 hours per day | High reliability | 20 GHS/m$^3$ or 0.40 GHS per 20-L bucket |
| 10 | Private in home | Chlorinated | 24 hours per day | Limited reliability | 20 GHS/m$^3$ or 0.40 GHS per 20-L bucket |
| 11 | Public handpump, 10 min | Unchlorinated | 24 hours per day | Limited reliability | 20 GHS/m$^3$ or 0.40 GHS per 20-L bucket |
| 12 | Public piped, 10 min | Chlorinated | 4 hours per day | Limited reliability | 20 GHS/m$^3$ or 0.40 GHS per 20-L bucket |
| 13 | Public handpump, 30 min | Chlorinated | 24 hours per day | High reliability | 10 GHS/m$^3$ or 0.20 GHS per 20-L bucket |
| 14 | Private on plot | Unchlorinated | 24 hours per day | High reliability | 10 GHS/m$^3$ or 0.20 GHS per 20-L bucket |
| 15 | Public handpump, 20 min | Chlorinated | 4 hours per day | High reliability | 10 GHS/m$^3$ or 0.20 GHS per 20-L bucket |
| 16 | Public piped, 10 min | Unchlorinated | 24 hours per day | Limited reliability | 10 GHS/m$^3$ or 0.20 GHS per 20-L bucket |
| 17 | Public handpump, 10 min | Chlorinated | 4 hours per day | Limited reliability | 10 GHS/m$^3$ or 0.20 GHS per 20-L bucket |
| 18 | Private in home | Unchlorinated | 4 hours per day | Limited reliability | 10 GHS/m$^3$ or 0.20 GHS per 20-L bucket |
| 19 | Public handpump, 10 min | Unchlorinated | 24 hours per day | High reliability | 2.5 GHS/m$^3$ or 0.05 GHS per 20-L bucket |
| 20 | Private in home | Unchlorinated | 4 hours per day | High reliability | 2.5 GHS/m$^3$ or 0.05 GHS per 20-L bucket |
| 21 | Public piped, 10 min | Unchlorinated | 4 hours per day | High reliability | 2.5 GHS/m$^3$ or 0.05 GHS per 20-L bucket |
| 22 | Private on plot | Chlorinated | 24 hours per day | Limited reliability | 2.5 GHS/m$^3$ or 0.05 GHS per 20-L bucket |
| 23 | Public handpump, 30 min | Chlorinated | 24 hours per day | Limited reliability | 2.5 GHS/m$^3$ or 0.05 GHS per 20-L bucket |
| 24 | Public handpump, 20 min | Chlorinated | 4 hours per day | Limited reliability | 2.5 GHS/m$^3$ or 0.05 GHS per 20-L bucket |

**Table S2.** Explanatory variables considered in the regression analysis focused on satisfaction with existing water services. Water source categories used by <25 respondents were removed. Wealth quintiles were derived using the DHS wealth index method. The final model was developed by sequentially removing variables with the largest p-values or variance inflation factors (VIF, a measure of collinearity), until all remaining variables had p<0.20 and VIF<3.

| Variable | Type | Possible values (with baseline level specified for categorical variables) | Included in final regression model |
|---|---|---|---|
| Region | Categorical | Upper West, Upper East, North East, Northern (baseline) | Yes |
| ***Household conditions*** | | | |
| Water source | Categorical | Piped water to public tap, mechanized borehole, protected dug well, unprotected dug well, surface water, borehole with handpump (baseline) | Yes |
| Household treatment | Binary | Yes, no (baseline) | No (p>0.20) |
| Available water is always sufficient for household | Binary | Yes, no (baseline) | Yes |
| Water collection time is within 30 minutes | Binary | Yes, no (baseline) | Yes |
| Water is accessible on premises (in home or on plot) | Binary | Yes, no (baseline) | Yes |
| Household pays for water | Binary | Yes, no (baseline) | Yes |
| Female respondent | Binary | Yes, no (baseline) | Yes |
| Respondent had no formal education | Binary | Yes, no (baseline) | No (p>0.20) |
| Household in bottom wealth quintile | Binary | Yes, no (baseline) | No (p>0.20) |
| Household in top wealth quintile | Binary | Yes, no (baseline) | No (p>0.20) |
| ***Community conditions*** | | | |
| Small town (vs. remote rural) | Binary | Yes, no (baseline) | No (p>0.20) |

| | | | |
|---|---|---|---|
| Community contains >100 households | Binary | Yes, no (baseline) | No (p>0.20) |
| Fraction of community in bottom wealth quintile | Percentage | 0 – 100% of households | No (p>0.20) |
| Fraction of community in top wealth quintile | Percentage | 0 – 100% of households | Yes |
| Fraction of household heads in community who are female | Percentage | 0 – 100% of households | No (p>0.20) |
| Fraction of household heads in community who had no formal education | Percentage | 0 – 100% of households | No (p>0.20) |
| Fraction of household heads in community who work in agriculture | Percentage | 0 – 100% of households | No (p>0.20) |
| Fraction of community paying for water | Percentage | 0 – 100% of households | Yes |
| Number of community water points | Integer | 1, 2, 3, … | No (p>0.20) |
| Any water points are functional | Binary | Yes, no (baseline) | No (p>0.20) |
| All water points are functional | Binary | Yes, no (baseline) | No (p>0.20) |
| Any water points are well-maintained | Binary | Yes, no (baseline) | No (p>0.20) |
| All water points are well-maintained | Binary | Yes, no (baseline) | No (p>0.20) |
| Any water points are well-maintained and clean | Binary | Yes, no (baseline) | No (p>0.20) |
| All water points are well-maintained and clean | Binary | Yes, no (baseline) | No (p>0.20) |
| Any water points are easily accessible | Binary | Yes, no (baseline) | No (p>0.20) |
| All water points are easily accessible | Binary | Yes, no (baseline) | No (p>0.20) |
| ***District-level activities*** | | | |

| | | | |
|---|---|---|---|
| District has used data for WASH planning | Binary | Yes, no (baseline) | No (p>0.20) |
| District has implemented water governance accountability mechanisms | Binary | Yes, no (baseline) | No (p>0.20) |
| District has an updated WASH investment plan | Binary | Yes, no (baseline) | No (p>0.20) |
| District has improved budget systems | Binary | Yes, no (baseline) | No (p>0.20) |
| District has conducted a WASH needs assessment | Binary | Yes, no (baseline) | No (p>0.20) |
| District has monitored piped water systems | Binary | Yes, no (baseline) | No (p>0.20) |
| District has documented water service providers | Binary | Yes, no (baseline) | No (p>0.20) |
| District has cited water service compliance issues | Binary | Yes, no (baseline) | No (VIF>5) |
| District has engaged with the private sector | Binary | Yes, no (baseline) | No (p>0.20) |

**Table S3.** Survey respondent characteristics from the baseline survey and additional DCE surveys conducted in rural small towns. To compare characteristics across different survey populations (DCE vs. non-DCE respondents in baseline, baseline vs. added respondents), we applied chi-squared tests in R, with p-values reported in the final two columns.

| Characteristic | All | All baseline respondents | Baseline DCE respondents only | Added small town DCE respondents | All DCE respondents | Baseline DCE vs. non-DCE (p-value) | Baseline vs. added (p-value) |
|---|---|---|---|---|---|---|---|
| Total respondents | 2172 | 1748 | 542 | 424 | 966 | | |
| Female respondents | 77.8% | 77.3% | 71.2% | 79.7% | 74.9% | <0.001 | 0.323 |
| Female household head | 11.7% | 12.7% | 11.1% | 7.8% | 9.6% | 0.195 | 0.006 |
| Household head had no formal education | 65.1% | 78.4% | 77.9% | 9.9% | 48.0% | 0.743 | <0.001 |
| Household head works in agriculture | 81.1% | 87.8% | 88.2% | 53.5% | 73.0% | 0.770 | <0.001 |
| Household size >7 members | 46.3% | 46.9% | 50.7% | 43.9% | 47.7% | 0.033 | 0.293 |
| Household in bottom wealth quintile | 20.0% | 24.6% | 19.6% | 1.2% | 11.5% | 0.001 | <0.001 |
| Household in top wealth quintile | 20.0% | 9.7% | 11.1% | 62.7% | 33.7% | 0.214 | <0.001 |
| Small town residents | 38.0% | 23.0% | 20.3% | 100.0% | 55.3% | 0.082 | <0.001 |
| *Water source* | | | | | | | |
| Piped water in home | 0.2% | 0.1% | 0.0% | 0.7% | 0.3% | 1.000 | 0.030 |
| Piped water on plot | 4.5% | 0.2% | 0.2% | 22.2% | 9.8% | 1.000 | <0.001 |
| Piped water to public tap | 7.5% | 3.0% | 3.3% | 25.9% | 13.3% | 0.748 | <0.001 |
| Water kiosk | 0.5% | 0.3% | 0.0% | 1.2% | 0.5% | 0.309 | 0.042 |
| Borehole with handpump | 56.0% | 62.1% | 59.4% | 30.9% | 46.9% | 0.138 | <0.001 |
| Mechanized borehole | 4.0% | 3.9% | 5.7% | 4.0% | 5.0% | 0.016 | 1.000 |
| Protected dug well | 2.3% | 1.8% | 1.3% | 4.5% | 2.7% | 0.350 | 0.002 |
| Unprotected dug well | 9.6% | 11.4% | 10.5% | 2.4% | 6.9% | 0.494 | <0.001 |
| Unprotected spring | 1.0% | 1.3% | 1.3% | 0.0% | 0.7% | 1.000 | 0.040 |
| Tanker truck | 0.1% | 0.1% | 0.4% | 0.2% | 0.3% | 0.178 | 1.000 |
| Surface water | 12.9% | 15.7% | 17.9% | 1.4% | 10.7% | 0.111 | <0.001 |
| Sachet or bottled water | 1.3% | 0.1% | 0.0% | 6.4% | 2.8% | 0.854 | <0.001 |
| Household water treatment | 7.7% | 8.4% | 9.2% | 4.7% | 7.2% | 0.465 | 0.014 |
| Household pays for water | 58.9% | 55.1% | 50.9% | 74.3% | 61.2% | 0.020 | <0.001 |
| Water collection time within 30 minutes | 72.3% | 70.1% | 74.4% | 81.1% | 77.3% | 0.012 | <0.001 |
| Water is always sufficient | 73.2% | 71.8% | 71.8% | 79.0% | 74.9% | 1.000 | 0.003 |
| Water is available on premises | 5.9% | 1.8% | 1.5% | 22.9% | 10.9% | 0.583 | <0.001 |
| *Inferred water service levels* | | | | | | | |
| At least basic water service | 56.8% | 51.7% | 53.7% | 78.1% | 64.4% | 0.277 | <0.001 |
| Safely managed water service | 4.1% | 1.1% | 0.7% | 16.7% | 7.8% | 0.488 | <0.001 |

#### SCENARIO 1: Would you be WTP

Pictures of tap water and chlorination (removed for publication)

**35 GHS/m3 or 70 pesewas per 20L bucket**

- Access to water from a **tap inside your dwelling**
- The water is **chlorinated**
- The water is typically available for **24 hours** per day
- Water service are interrupted no more than 1-2 days per month
- Breakdowns never last more than 24 hours per day because someone performs regular maintenance

Pictures of currency to illustrate amount (removed for publication)

= **35 GHS/M3**

#### SCENARIO 6: Would you be WTP

Pictures of handpump and women walking while carrying water (removed for publication)

**35 GHS/m3 or 70 pesewas per 20L bucket**

- Access to water from a **public hand pump** where it takes **30 minutes** to go, collect water, and return home
- The water is **not chlorinated**
- The water is typically available for **4 set hours** per day
- Water service is interrupted 2 days per week
- Breakdowns can last more than 24 hours because maintenance only happens over a breakdown.

Pictures of currency to illustrate amount (removed for publication)

= **35 GHS/M3**

**Figure S1.** Examples of informational handouts used to explain scenarios to respondents during discrete choice experiments. These handouts show Scenario 1 (top, representing the highest overall service level) and Scenario 6 (bottom, representing the lowest service level). Note: The pictures associated with these scenarios have been removed per medRxiv requirements, and are available from the corresponding author upon request.

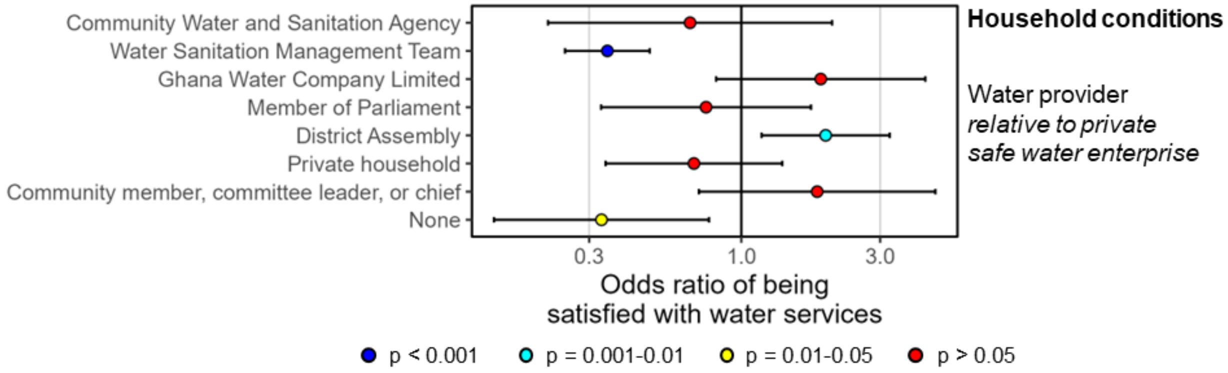


**Figure S2.** Associations between household satisfaction with water services and water service provider, based on logistic regression. For the purpose of this regression, the four-level Likert scale was consolidated into two levels: satisfied (including "satisfied" and "very satisfied") and unsatisfied (including "somewhat unsatisfied" and "very unsatisfied"). The regression model included adjusted standard errors to account for community clustering. Variables with odds ratios above 1 are positively associated with satisfaction, while odds ratios below 1 are negatively associated. Error bars represent 95% confidence intervals. The reference level for the water service provider categorical variable was private safe water enterprises, which included water points provided or managed by non-governmental organizations (NGOs) and was the most common type.

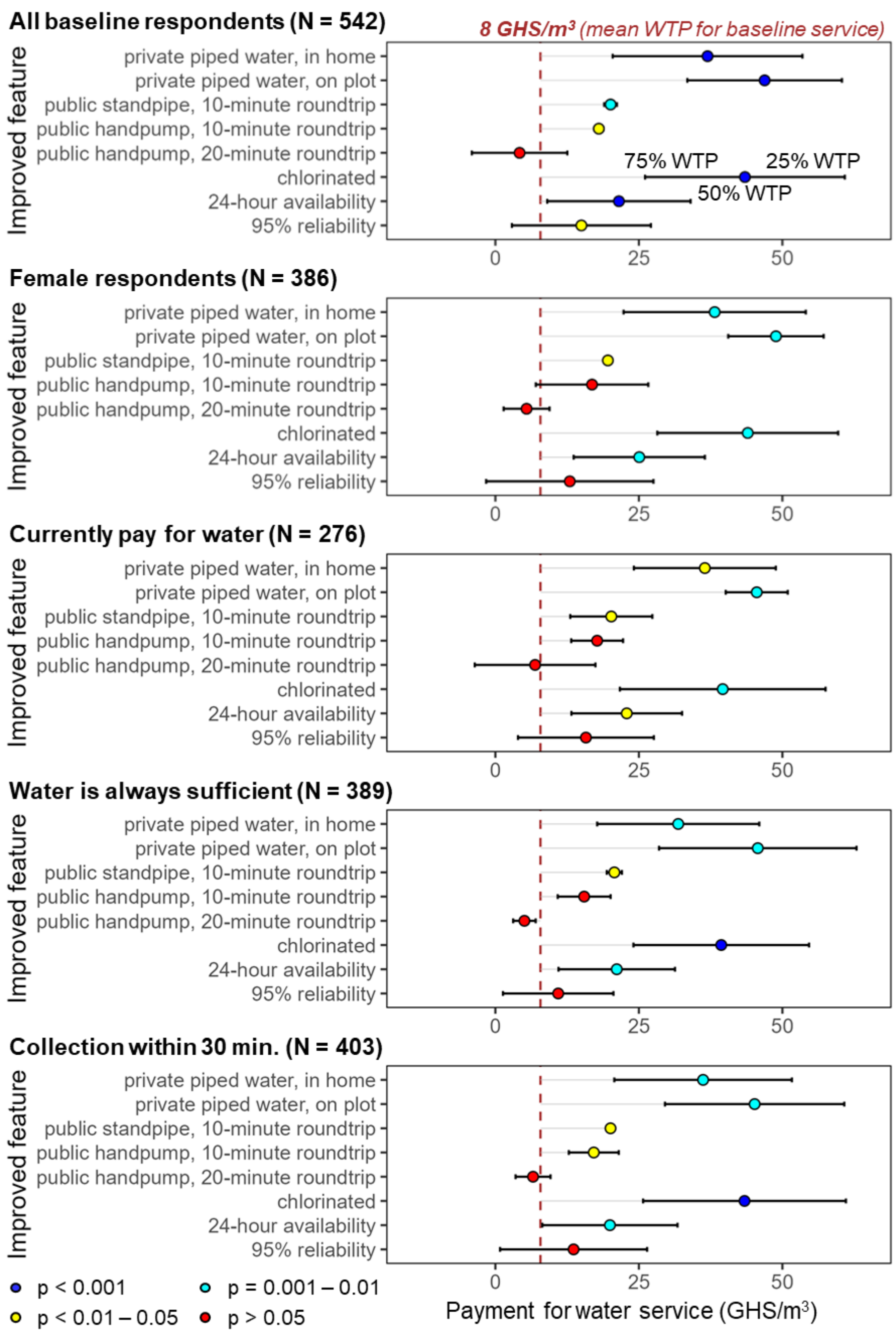


**Figure S3.** Estimated willingness-to-pay for service improvements relative to a baseline level of service (defined as access to an unchlorinated public handpump that is a 30-minute roundtrip from the household, with water available 4 hours per day and breakdowns 30% of the time), based on discrete choice experiments (DCE) among various subsets of the baseline survey population. The top graph provides DCE results for the full population (equivalent to Figure 4 in the main text) for comparison. Each graph below it presents the results of mixed logit models using subsets of the baseline survey population, focusing especially on characteristics associated with satisfaction with existing water services. The dashed red line shows mean WTP for the reference service level, as stated by respondents in response to an open-ended question. Points represent estimates of mean WTP when an improved feature is added, and error bars represent the first and third quartiles of the modeled WTP distribution (modeled as a normal distribution based on individual preferences). All models produced similar trends, with estimates of mean WTP for each feature remaining very close to those from the original model. We interpret these findings to mean that personal preferences, rather than broader demographic or service characteristics, largely determined variations in estimates of individual WTP.

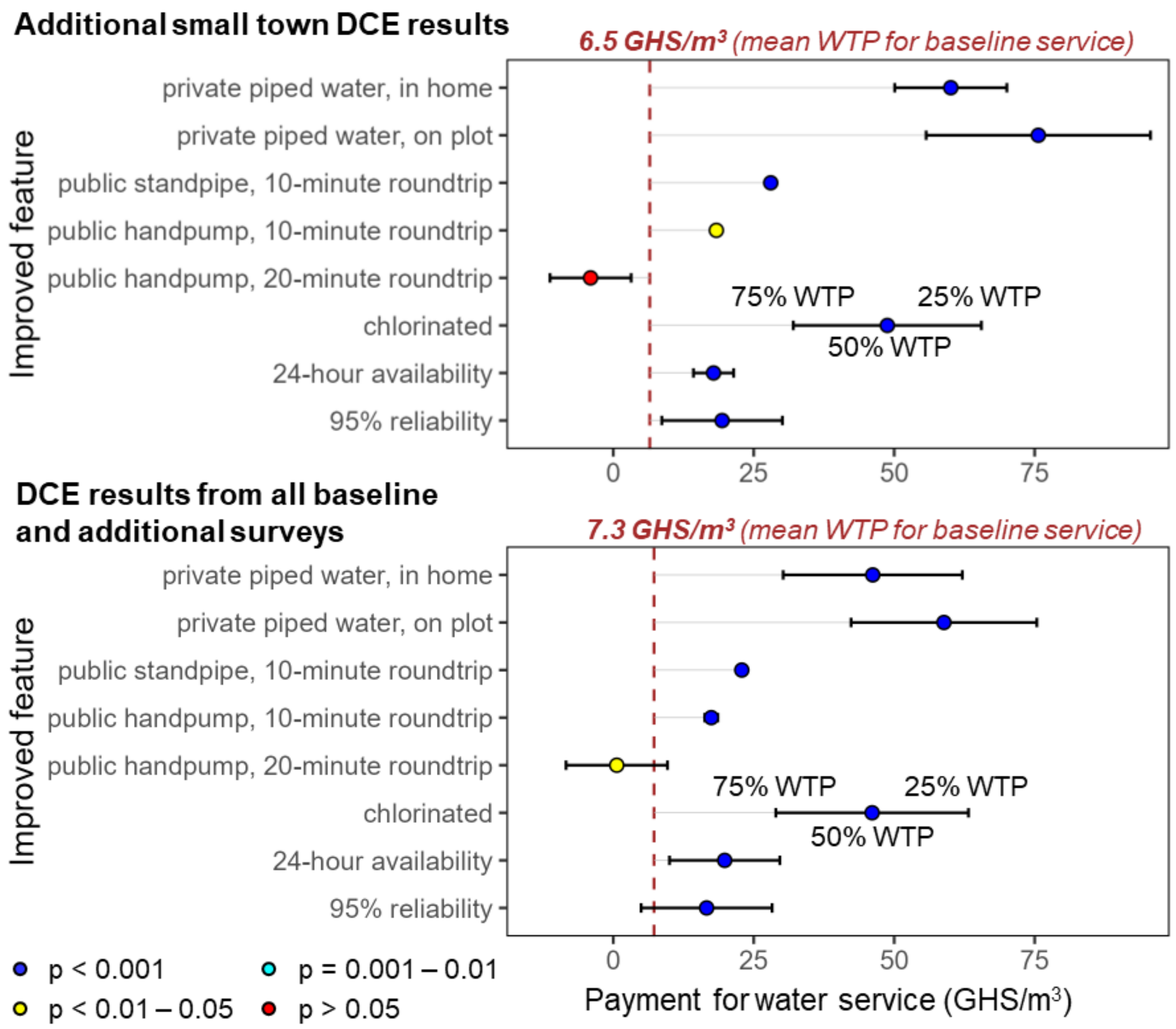


**Figure S4.** Estimated willingness-to-pay for service improvements relative to a baseline level of service (defined as access to an unchlorinated public handpump that is a 30-minute roundtrip from the household, with water available 4 hours per day and breakdowns 30% of the time), based on discrete choice experiments (DCE) among additional small town survey participants (top, N=424) and among all DCE participants across the baseline and additional surveys (bottom, N=966). Figure 4 in the main text shows results from baseline DCE participants only (N=542). The dashed red lines show mean WTP for the baseline service level, as stated by respondents in response to an open-ended question. Points represent estimates of mean WTP when an improved feature is added, and error bars represent the first and third quartiles of the modeled WTP distribution (modeled as a normal distribution based on individual preferences). Accordingly, results where error bars cross the baseline mean (e.g., 95% reliability) suggest that at least 25% of respondents may not be willing to pay more for these features, even if a majority do value them more highly than the baseline level. Note that error bars for the public handpump are not visible because their values are extremely close to the point estimate.
